# Experimental and computational evaluation of knee implant wear and creep under in vivo and ISO boundary conditions

**DOI:** 10.1101/2023.05.09.23289712

**Authors:** Michael J. Dreyer, Seyyed Hamed Hosseini Nasab, Philippe Favre, Fabian Amstad, Rowena Crockett, William R. Taylor, Bernhard Weisse

**Affiliations:** Laboratory for Movement Biomechanics, Institute for Biomechanics, ETH Zürich, Switzerland; Laboratory for Mechanical Systems Engineering, Empa, Dübendorf, Switzerland; Zimmer Biomet, Zug, Switzerland; Laboratory for Surface Science and Coating Technologies, Empa, Dübendorf, Switzerland

**Keywords:** Implant, knee, wear, creep, model, simulation

## Abstract

Experimental knee implant wear testing according to ISO 14243 is a standard procedure, but it inherently possesses limitations for preclinical evaluations due to extended testing periods and costly infrastructure. In an effort to overcome these limitations, we hereby develop and experimentally validate a finite element (FE) based algorithm, including a novel cross-shear and contact pressure dependent wear and creep model, and apply it towards understanding the sensitivity of wear outcomes to the applied boundary conditions.

Specifically, we investigated the application of in vivo data for level walking from the publicly available “Stan” dataset, which contains single representative tibiofemoral loads and kinematics derived from in vivo measurements of six subjects, and compared wear outcomes against those obtained using the ISO standard boundary conditions. To provide validation of the numerical models, this comparison was reproduced experimentally on a six-station knee wear simulator over 5 million cycles, testing the same implant Stan’s data was obtained from.

Experimental implementation of Stan’s boundary conditions in displacement control resulted in approximately three times higher wear rates (4.4 vs. 1.6 mm^3^ per million cycles) and a more anterior wear pattern compared to the ISO standard in force control. While a force-controlled ISO FE model was unable to reproduce the bench test kinematics, and thus wear rate, displacement-controlled FE models accurately predicted the laboratory wear tests for both ISO and Stan boundary conditions. The credibility of the in silico wear and creep model was further established per the ASME V&V-40 standard. The model is thus suitable for supporting future patient specific models and development of novel implant designs.

## 1. Introduction

Longevity of knee implants is a major concern for the two thirds of total knee arthroplasty (TKA) patients who are less than 65 years old (Kurtz et al., 2009). Today, long term failure of knee implants due to wear of the polyethylene (PE) inlay (Kim et al., 2014) or related to aseptic loosening (Oparaugo et al., 2001; Pitta et al., 2018) still occurs, despite improvements in implant designs and material such as PE crosslinking. Efforts to comprehensively investigate and increase the long-term wear resistance of knee implants, however, are somewhat constrained by limited accessibility and viability of current preclinical wear testing methods.

Knee implant wear is typically evaluated by means of laboratory wear tests. However, experimental wear testing is extremely time-consuming and costly: A test running for five million cycles at 1 Hz takes approximately four months to complete (Haider, 2009). Thus, experimental implant wear testing is not a practicable tool to compare more than a few conditions or designs at a time. Moreover, variability in the outcome wear measures can be considerable and repeatability may be challenging to achieve (Abdelgaied et al., 2011).

To provide a viable alternative, computational wear simulations, mostly based on deformable finite element (FE) models, have been developed (Abdelgaied et al., 2018; Fregly et al., 2005; Mell et al., 2018). Such computational models have proven to strongly complement experimental testing by being orders of magnitude faster and not requiring dedicated personnel and infrastructure (Taylor and Prendergast, 2015). This allows evaluating the influence of parameters such as implant design, implant positioning, or loading conditions on wear, each taken individually or simultaneously in probabilistic studies (Pal et al., 2008). However, for such models to be useful, their credibility must first be established (Viceconti et al., 2009).

When mechanical and in silico wear simulations aim to predict in vivo wear, applied loads and kinematics should be representative of in vivo conditions. While such data has historically been scarce, in vivo implant loads and kinematics have now been made publicly available as part of the CAMS-Knee dataset (Taylor et al., 2017). More recently, the data from the 6 CAMS-Knee subjects were standardized into the single averaged “Stan” dataset, and thus made accessible for mechanical wear simulation (Dreyer et al., 2022b). Interestingly, the commonly used ISO 14243-1 standard loads and kinematics (ISO, 2009), which were calculated from simplified models, were shown to differ from the Stan loads and kinematics measured in vivo for level walking.

In this exploratory study, we firstly aimed to develop and validate an advanced computational wear and creep model for predictions of how patient- and implant-specific factors impact PE inlay wear. To this end, a cross-shear and contact-pressure dependent wear model was combined with a novel creep prediction method. The necessary input material data was obtained from fully independent experimental studies. Secondly, the first comparison of wear resulting from the application of Stan’s loads and kinematics to wear induced from the ISO standard boundary conditions (BCs) was performed using both computational simulation and experimental testing of wear.

## 2. Methods

### Wear test

Related to the first aim of the study, the main purpose of the experimental wear test was to validate the computational wear and creep model (see below). Additionally, it served to compare the effect of the two different BCs on wear for the second aim of this study. For consistency, all experiments and computational simulations were performed on the same ultra-congruent cruciate-sacrificing TKA implant (Innex® FIXUC, Zimmer Biomet, Switzerland), which was also implanted in the patients involved in the CAMS-Knee and Stan investigations.

Implant components were tested on a six-station knee simulator (AMTI, Watertown, USA), which allowed control of femoral flexion angle and anterior-posterior (AP) force or translation, as well as tibial axial force and internal-external (IE) moment or rotation (Figure 1). The tibial component was fixed to have a posterior slope of 6° according to the manufacturer’s surgical technique.

**Figure 1.**
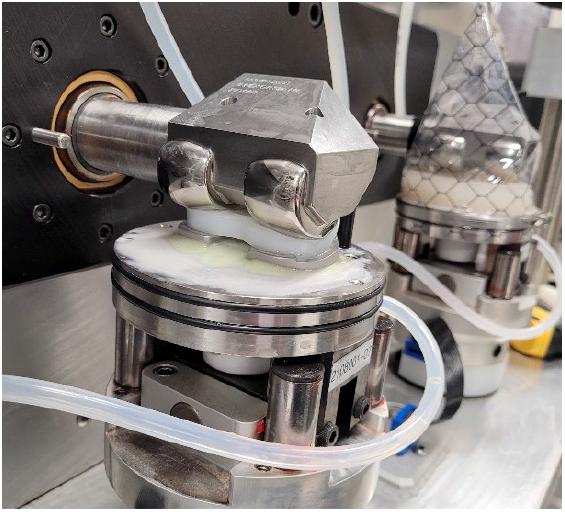
Wear test specimens consisting of PE inlay, tibial, and femoral components fixed in the bench test setup in dry (foreground) and sealed with lubricant (background) state.

The test lubricant was bovine calf serum (Hyclone™ Calf Serum, Cytvia, USA) diluted to a protein concentration of 20 g/L (ISO 12443-1, ISO, 2009). Additionally, 7.4 g/L ethylenediaminetetraacetic acid disodium salt dihydrate (Fisher Scientific, USA) and 2.0 g/L sodium azide (Fisher Scientific, USA) were added to hinder bacterial growth and build-up of calcium phosphate on the implant surfaces according to ASTM F732 – 17 (ASTM International, 2017a). The specimens were pre-soaked in this lubricant for 12 weeks prior to testing.

In the bench test setup (Figure 1), the ISO BCs as well as Stan’s kinematics for level walking and the associated CAMS-HIGH100 loads (Dreyer et al., 2022b) were applied to the implants (Table 1). Stan’s data were applied in displacement-control (DC) to closely reproduce the in vivo contact mechanics in the bench test and models. For consistency, Stan’s conditions would ideally have been compared to the DC ISO 14243-3:2014 standard. However, preliminary FE simulation of the DC ISO standard showed excessive posterior edge loading, also observed in other studies (Abdelgaied et al., 2022; Zhang et al., 2019), which led to dislocation of the implant. Therefore, the force-controlled (FC) ISO 14243-1:2009 standard was used as a comparison for Stan’s loads and kinematics.

**Table 1.**
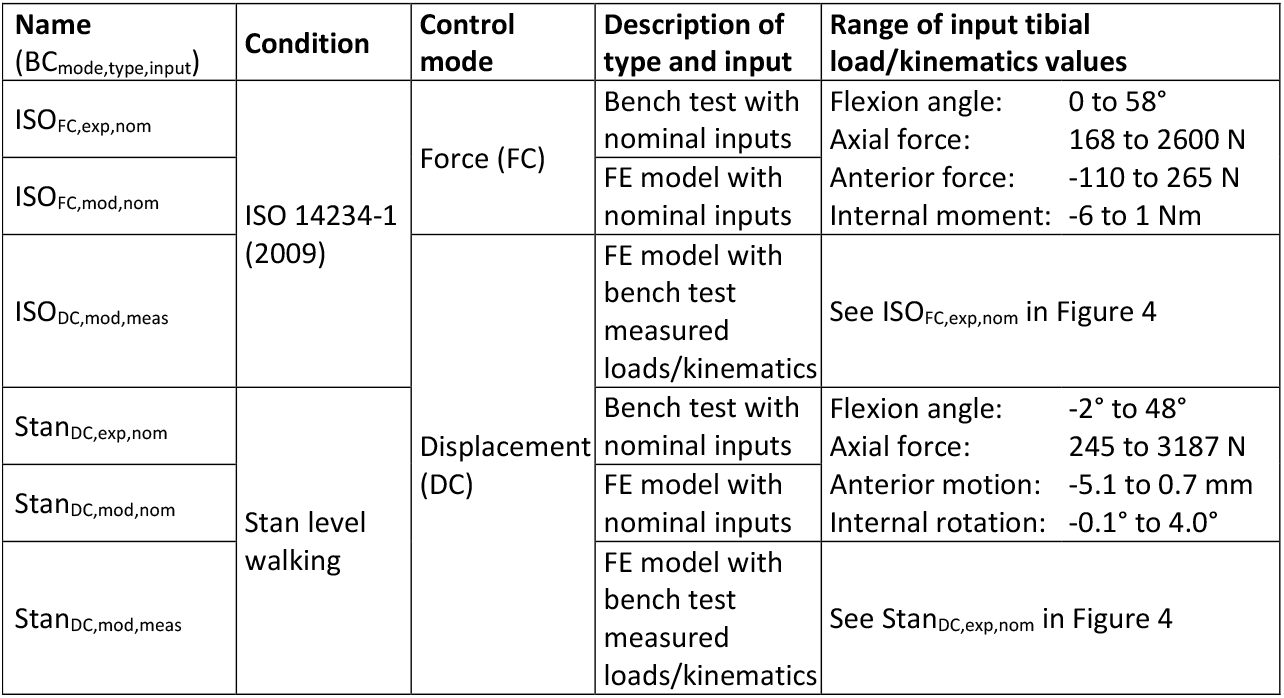
Overview of experimental and modelling boundary conditions. Note that flexion angle and axial force are driven in displacement and force control, respectively, for all tests regardless of the stated control mode, for consistency with ISO standards.

One group of three specimens was subjected to the ISO FC BCs and one group of three specimens was subjected to Stan’s kinematics in DC mode (ISO_FC,exp,nom_ and Stan_DC,exp,nom_ in Table 1, respectively). The ISO AP force, IE moment, and flexion angle were applied as per the FC standard (ISO 12443-1, ISO, 2009). Stan’s CAMS-HIGH100 loads and kinematics were transformed to the ISO coordinate system and AP and IE kinematics (DC mode) and axial force were applied consistent with the DC ISO standard (ISO 12443-3, ISO, 2014). For each group, two additional soak-control specimens were submerged in lubricant and subjected to the same axial load profile as the wear specimens, but without any other loads or motion, to correct for PE weight changes due to fluid uptake.

The test was performed at 1.1Hz for 5 million cycles (MC). Wear was measured gravimetrically at 500’000 cycles and afterwards at every full MC until test completion. Subsequently, the volumetric wear rate of each PE inlay was calculated from the slope of the regression line fitted to the wear volume over number of cycles, assuming a PE density of 0.935 g/cm^3^ (Saikko, 2017). The effective loads and kinematics applied by the testing machine to the implants were recorded every 20’000 cycles to allow the average applied loads and kinematics to be calculated.

Before and after the test, the three-dimensional inlay geometry was measured using a structured light 3D scanner (Pro S3, HP Inc., USA) with a resolution of ∼50 μm. The untested and tested 3D geometries of each specimen were aligned using an iterative closest point algorithm and the change in surface geometry due to wear and creep was plotted using custom Python scripts.

### Finite element model

Finite element (FE) models of the experimental test setup (Figure 2) were created in Abaqus/Standard 6.21 (Dassault Systèmes, USA). These models consisted of the PE inlay and the femoral component, with tibio-femoral contact defined by a coefficient of friction of 0.04 (Godest et al., 2002). The tibial component was not modelled, as the predicted backside wear on the fixed inlay would be minimal (O’Brien et al., 2013). To enable faster convergence, automatic tangential contact damping was activated, but scaled down by a factor of 0.0001 after confirming a negligible impact on model outputs. The inlay was assigned elastic-plastic material properties calibrated by the manufacturer to material characterization tests on the PE used in the Innex implant. Element size was chosen based on a convergence study on contact pressure and wear, reaching a change in output <2% between two successive mesh refinements. The inlay was assigned a general element size of 2.5 mm, with 0.9 mm elements on the contact surfaces, resulting in 41’833 quadratic tetrahedral elements. The femoral component was modelled as a rigid shell (Carr and Goswami, 2009) with an element size of 0.5 mm on the contact surfaces and approximately 2 mm on the sides, for a total 22’764 linear quadrilateral and triangular elements. The testing machine’s fixtures were represented by rigid connector elements (Figure 2) to which the Stan and ISO input loads and kinematics were then applied, resulting in the ISO_FC,mod,nom_ and Stan_DC,mod,nom_ models (Table 1). Each boundary condition motion cycle was split into 200 time intervals, based on a temporal convergence study considering volumetric wear, leading to a difference of <0.2% between two interval sizes.

**Figure 2.**
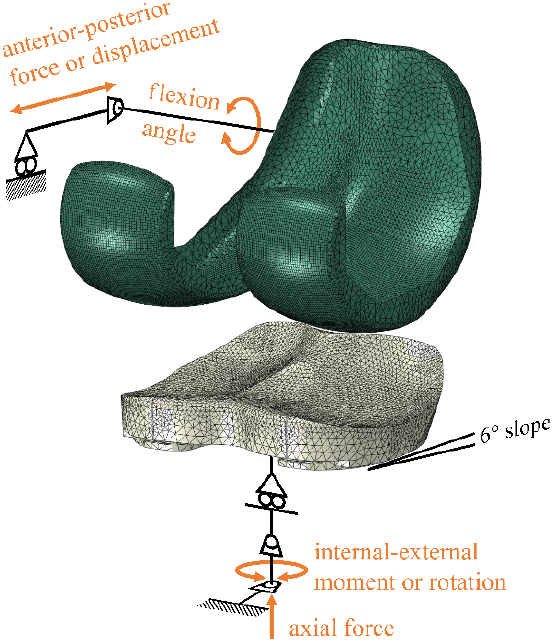
Exploded view of the finite-element model, consisting of the inlay (beige) and femoral (green) components and the rigid connector elements representing the wear simulator fixtures.

For the same input loads or kinematics, slightly different resultant contact loads and kinematics could be expected between the knee simulator, which is affected by inertia, tolerances, and control system delays, and the FE model, which is affected by simplifications and idealized component geometries. This is especially the case for FC mode, where deviations of several millimetres and degrees commonly occur (Abdelgaied et al., 2022; Bauer et al., 2021; Knight et al., 2007). To evaluate the influence of these different contact mechanics on wear and creep prediction, two additional simulations were run. Specifically, the average kinematics and axial forces of the ISO and Stan groups measured on the testing machine were applied to the corresponding FE models in DC mode (ISO_DC,mod,meas_ and Stan_DC,mod,meas_ in Table 1). In this manner, the wear rates of the FE model and experiment were evaluated under identical contact kinematics.

### Wear and Creep Prediction Algorithm

Implant wear is known to depend on contact mechanics, but contact mechanics progressively change if the surface geometry is altered by wear or creep (Zhang et al., 2017). To ensure appropriate modelling of wear (Knight et al., 2007; Zhao et al., 2008), this interdependence was reproduced iteratively in our wear and creep prediction algorithm (named “WearPy”), a custom Python code that directly interacts with Abaqus (Figure 3).

**Figure 3.**
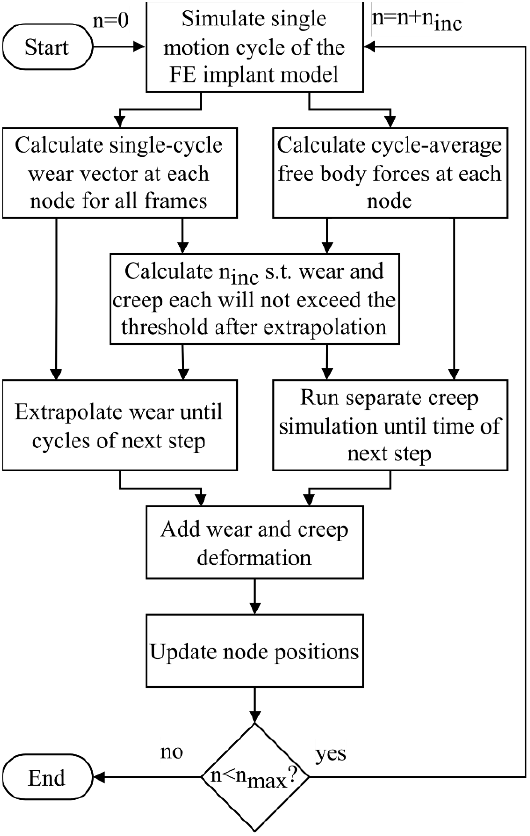
Flowchart of the “WearPy” implant wear and creep prediction pipeline. Here, n represents the current number of cycles in the analysis, which is iterative with steps of fewer cycles n_inc_ and stops when the maximum number of cycles n_max_ is reached.

WearPy divides the total number of cycles n_max_ (5 MC here) into steps of fewer cycles n_inc_. Each step consists of the solution of the FE model described above for a single motion cycle, the calculation of wear and surface loads from the results, and the extrapolation of the wear and simulation of the creep over n_inc_ cycles until the next step. The removal of material due to wear and the deformation due to creep are modeled by updating the surface nodal positions. A preliminary convergence study on a pin-on-disk model showed that a change in surface geometry of up to 0.01 mm would not significantly change the contact mechanics. Thus, to ensure a smooth progression of surface deformation (Zhao et al., 2008), WearPy automatically chose the largest possible number of cycles per step (n_inc_) such that the larger of wear and creep caused surface deformations of exactly 0.01 mm and the smaller of wear and creep consequently caused <0.01 mm of deformation. After calculating a step wear and creep for the chosen number of cycles, the inlay mesh was updated, and the whole procedure was repeated until n_max_=5 MC was reached.

### Wear model

To model the critical influence of cross-shear and contact-pressure on wear (Baykal et al., 2014), the local wear depth δ at each node was calculated using a modified version of Archard’s Law:

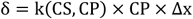

Here, Δx is the sliding distance, and k(CS,CP), measured in mm^3^N^-1^m^-1^, is the wear factor as a function of cross-shear ratio CS as calculated by Goreham-Voss et al. (2010) and contact-pressure CP. The wear factor k(CS,CP) was defined as:

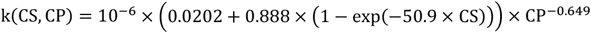

This empirical expression was derived from comprehensive pin-on-disk wear tests performed on the same PE material from which the inlays in this study are made (Dreyer et al., 2022a).

### Creep model

To improve the accuracy of the contact-pressure dependent wear model, it is necessary to include surface deformations due to creep in the model (Quinci et al., 2014). Thus, a model for dynamic compressive creep of PE from the literature (Lee et al., 2003) was adapted, assuming 50% of creep deformation would be recovered (Lee and Pienkowski, 1998) during the test interruptions to measure gravimetric wear as well as after the test. The formula to calculate the creep strain ε_creep_ based on the von-Mises stress σ_VM_ and the time in minutes, t_minutes_, was defined as:

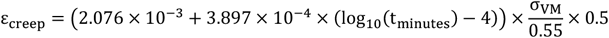

This equation was implemented into the Abaqus “CREEP” user-subroutine. As part of the wear prediction algorithm, a separate creep analysis was automatically performed to determine the geometrical changes that occur over the number of cycles n_inc_ between two steps. During this creep analysis, the average over time of all the free-body-forces from contact and boundary conditions acting on each node of the inlay during the motion cycle was extracted from the solution of the FE model described above and then, in a separate creep simulation, applied as a static load to each node of the inlay. Thus, creep deformation was calculated for the whole inlay. To our knowledge, this is the first study to consider creep of a whole knee implant component, as other studies only modelled creep in a local contact area (Abdelgaied et al., 2011; Fregly et al., 2005; Willing and Kim, 2009; Zhao et al., 2008).

### Verification and Validation

To assess the credibility of the modelling approach and WearPy, verification and validation was performed according to ASME V&V 40-2018 (ASME, 2018) and regulatory (FDA, 2021) guidelines (see supplementary material). To this end, the validation included a comparison of the FE models and the experimental test comparator for the ISO and Stan conditions. A hypothetical context of use was defined, where the wear would be predicted during development of a new knee implant to identify the worst-case configuration for experimental wear testing. Finally, each of the credibility factors was independently assessed relative to the model risk associated with using the model here to support comparative evaluation of TKA designs.

## 3. Results

Three different sets of results were obtained: First the knee simulator test, second the corresponding FE models with the same input data, and third the FE models with kinematics input that was directly measured in the knee simulator test. For each of the three sets, Stan’s in vivo condition and for the standard ISO condition are reported.

### Joint loads and kinematics

#### Stan BC vs. ISO BC

Comparing the outputs, the test and models with Stan’s BCs exhibited higher peak axial forces (∼3187 N vs. ∼2600 N) and peak external moments (9.6 to 20.5 Nm vs. 1.9 to 7.1 Nm) than with the ISO BCs (Figure 4). For the kinematics, however, Stan’s BCs resulted in lower peak flexion angles (∼49° vs ∼58°), tibial internal rotation angles (∼3.9° vs. 7.5° to 10.3°), and tibial anterior translation (0.6 to 0.7 mm vs. 4.1 to 5.5 mm) than the ISO BCs.

**Figure 4.**
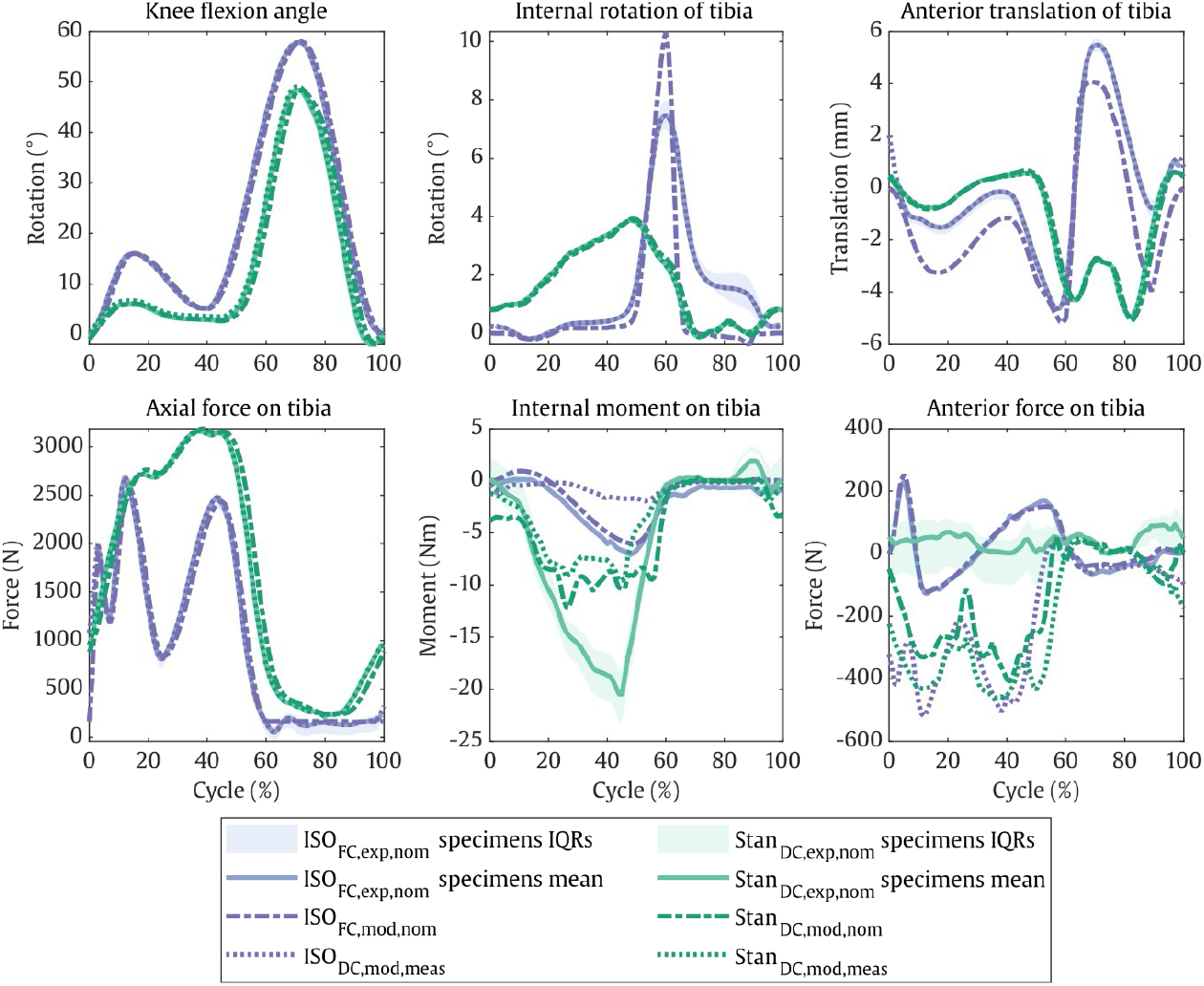
Resulting joint loads and kinematics measured for the specimens on the simulator (mean over all test intervals and specimens and interquartile ranges (IQRs) of each specimen), simulated using finite-element analysis with the same inputs (ISO_FC,exp,nom_ and Stan_DC,exp,nom_), and simulated using the experimentally measured kinematics as input to the FE model (ISO_DC,exp,meas_ and Stan_DC,exp,_). Forces and moments are expressed as external loads acting on the articulating surface of the tibial inlay.

#### ISO BC: Experiment vs. model

For the ISO_FC,exp,nom_ test and ISO_FC,mod,nom_ model with the same inputs, there was no more than 23 N difference in peak output AP force and 1 Nm in IE moment (Figure 4). The kinematics peaks, however, deviated by up to 1.4 mm in the AP and up to 2.9° in the IE directions, especially during swing phase. Moreover, in the ISO_FC,exp,nom_ test, the peak internal rotation and anterior translation values varied by up to 2.5° and 1 mm over the course of the test. When the test’s experimentally measured kinematics were applied to the ISO_DC,mod,meas_ model, tibial loads differed from the FC ISO standard input loads.

Specifically, AP contact forces acted only posteriorly and exceeded 500 N, compared to 230 N in the anterior and 130 N in the posterior directions for the ISO_FC,exp,nom_ test and ISO_FC,mod,nom_ model with nominal ISO inputs. External moments of the ISO_DC,mod,meas_ model were low, however, at only 1.9 Nm compared to 7.1 Nm in the wear test.

#### Stan BC: Experiment vs. model

For the Stan DC test and two FE models, the output kinematics were in close agreement with differences in peak AP translation and internal rotation of less than 0.2 mm and 0.1°. In contrast, load deviations between the experiment and the DC models were observed. Mostly anterior tibial forces of up to 93.5 N were observed experimentally, while both the Stan_DC,mod,nom_ and Stan_DC,mod,meas_ models predicted mostly posterior forces of up to 408 N and 461 N, respectively (Figure 4). Again, the modelled external moments of up to 12.2 Nm were lower compared to up to 20.4 Nm measured in the wear test. Moreover, in the Stan_DC,exp,nom_ test, the peak internal moments and anterior forces varied by up to 8 Nm and 200 N over the course of the test.

### Wear

The experimentally measured linear volumetric wear rate from 0.5–5.0 MC was 1.3–1.9 mm^3^/MC for the ISO_FC,exp,nom_ group and a more than two-and-a-half times higher 3.5–4.9 mm^3^/MC for the Stan_DC,exp,nom_ group (Figure 5). The Stan_DC,mod,nom_ model predicted a wear rate of 4.3 mm^3^/MC, falling well within the experimental range for these BCs. This was not the case for the ISO_FC,mod,nom_ model, which predicted a wear rate of 4.8 mm^3^/MC, being about three times higher than the corresponding experimentally obtained values. In contrast, both the ISO_DC,mod,meas_ and Stan_DC,mod,meas_ models driven by the experimental kinematics predicted wear rates that fell within the experimental ranges at 1.6 and 3.5 mm^3^/MC, respectively (Figure 5).

**Figure 5.**
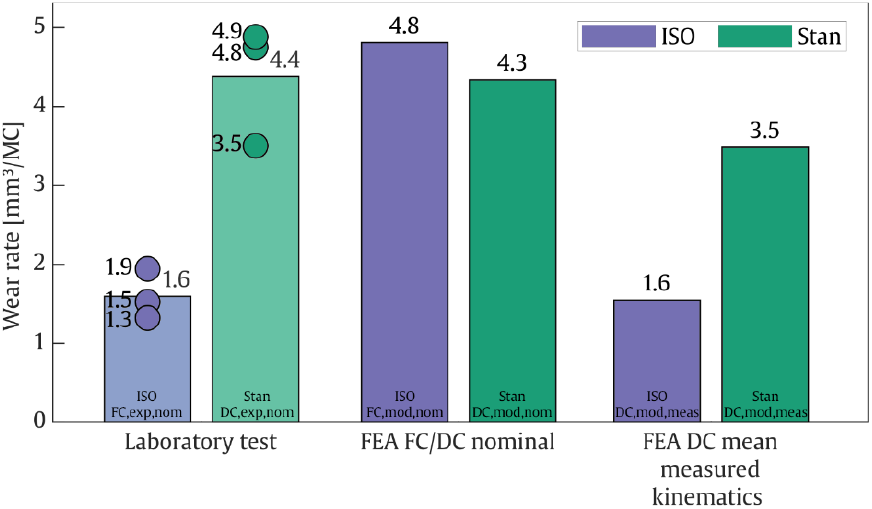
Wear rates measured for the specimens on the simulator (Laboratory test), simulated using finite-element analysis with the same inputs (FEA FC/DC nominal), and simulated using the experimentally measured kinematics as inputs to the FE model (FEA DC mean measured kinematics).

### Surface Deformation

The ISO_FC,exp,nom_ test induced clear surface deformation to the medial and posterior facets of the inlay, even though the overall wear rate was lower than for Stan_DC,exp,nom_ (Figure 6). The ISO_DC,mod,meas_ model with the same kinematics showed similarly posterior surface deformation, while the ISO_FC,mod,nom_ model showed posterolateral and anteromedial surface deformation. For the DC Stan condition, there was posterolateral and posterolateral surface deformation in the laboratory test and both models. Overall, the combined surface deformation induced by wear and creep in the FE simulations showed qualitative visual agreement with the 3D scan measurements on the corresponding physical test specimens.

**Figure 6.**
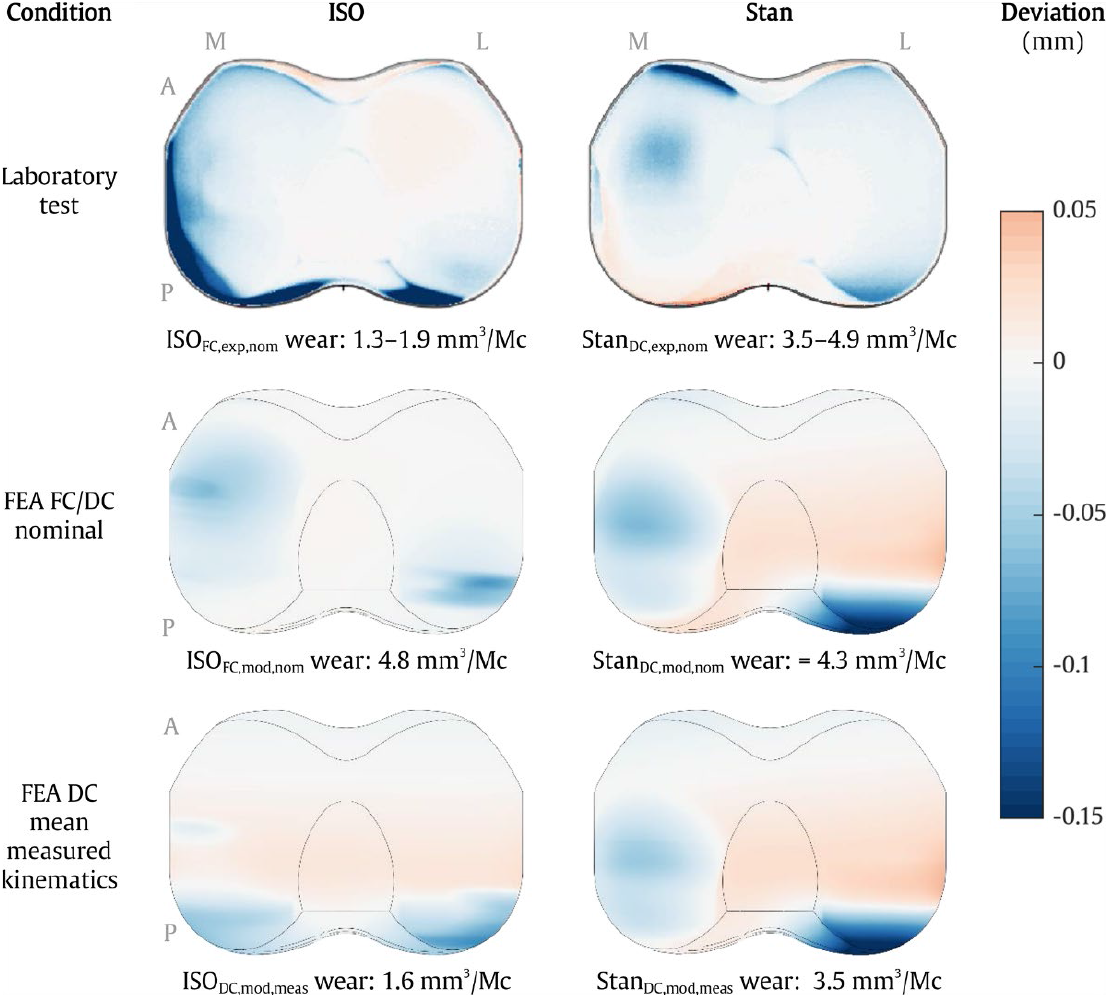
Surface deviation in the axial direction caused by wear and creep after 5 MC for the four models and the test specimens. Each test specimen plot represents the mean deviation of the three corresponding specimens.

### Verification and Validation

The credibility of the verification and validation activities was evaluated in accordance with standardized (ASME, 2018) and regulatory (FDA, 2021) guidance on model credibility (see supplementary material). Implementation of the scheme from the FDA guidance had practical challenges associated with mapping the proposed five-level gradation scheme for risk (the x-axis in Supplementary Figure S1) to credibility factors that have either three or four gradations (the y-axis). Nevertheless, the resulting evaluation provided a visual and easily interpretable overview of the credibility of the model. The conclusion from this assessment was that the modeling approach here is credible for use in support of low to medium model risk applications.

## 4. Discussion

Laboratory based testing of knee implant wear based on the currently established ISO boundary conditions is expensive and time consuming. Therefore, laboratory wear testing is rarely feasible for larger scale investigations into the effect of implant design, patient or surgical factors, or activity on implant wear. In an effort to address this challenge, the efficacy of FE simulation coupled with advanced PE wear and creep models as an alternative method of wear quantification was demonstrated here. Systematic model verification and validation was carried out, and the first comparison of the recently published standardized tibiofemoral implant loads and kinematics (“Stan”, Dreyer et al., 2022b) against the ISO boundary condition was performed. To our knowledge, this is also the first study investigating wear of the Innex knee implant by means of laboratory testing or computational modelling, hence providing quantitative evidence supporting the widely used CAMS-Knee datasets (Taylor et al., 2017).

The main finding was that Stan’s loads and kinematics resulted in higher wear rates and different surface deformation patterns compared to the ISO FC standard. Moreover, the FE wear model was able to accurately predict experimentally obtained wear, but was shown to be sensitive to inaccurate calculation of the joint kinematics. The experimental wear rates of 1.3–1.9 mm^3^/MC for the ISO_FC,exp,nom_ test and 3.5–4.9 mm^3^/MC for the Stan_DC,exp,nom_ test were lower than expected, however, compared to 5–40 mm^3^/MC for various other knee implant models with inlays made from conventional non highly crosslinked PE (Okazaki et al., 2019). The wear occurring more anteriorly on the medial than on the lateral condyle is consistent with earlier investigations of tibiofemoral contact locations for the CAMS-Knee data on the same implant model (Trepczynski et al., 2019) and of wear for similar in vivo data on another implant design (Wang et al., 2019).

The low wear rate of the ISO_FC,exp,nom_ experiment can be explained by the contact occurring mainly on the posterior inlay edge. This is evident from the visible deformation for both the ISO_DC,mod,meas_ model and the 3D scans (Figure 6). The posterior contact led to a small contact area, resulting in little overall implant wear. In comparison, the ISO_FC,mod,nom_ model with nominal inputs predicted contact to occur roughly 2 mm more anteriorly on the inlay (Figure 4, top right). This resulted in a larger contact area (Figure 6, middle left), more relative sliding and less rolling, and almost three times more wear (Figure 5), as a larger contact area can increase wear even with the same load applied (Abdelgaied et al., 2018; Kang et al., 2009; Saikko, 2006). The large wear rate mismatch between the ISO_FC,mod,nom_ model and the corresponding ISO_FC,exp,nom_ experiment could therefore be due to limited representation of the simulator machine’s inertial, friction, and control properties in the FC FE model, limiting its predictive capabilities with respect to joint kinematics. This is a common limitation of FC computational models (Abdelgaied et al., 2022; Bauer et al., 2021; Knight et al., 2007) and does not necessarily indicate poor modelling of the wear mechanism itself.

Wear was accurately predicted by our algorithm. The DC models driven by nominal (Stan_DC,mod,nom_) and measured (ISO_DC,mod,meas_ and Stan_DC,mod,meas_) kinematics all predicted wear rates within the range of experimentally measured values. The experimental surface wear patterns were more spread out than the model predictions and showed edge deformations. This is likely due to the observed variability in kinematics over the course of the test and for the ISO_DC,exp,nom_ due to an unwanted motion that occurred once after a test restart and deformed the inlay edges. However, the surface deformation patterns still qualitatively matched the three DC models. All this was achieved without tuning the models’ underlying material data to the validation experiments in any way, rather the material data was obtained from separate experiments. Wear volume and pattern were inaccurately predicted only for the ISO_FC,mod,nom_ model, where the joint kinematics differed most compared to the experiment. This shows that knee implant wear can be predicted accurately if the underlying joint model is able to reliably reproduce the real-world joint contact mechanics, but may be inaccurate if not, highlighting the importance of exact in vivo measurements of joint kinematics for patient-specific models.

Specifically, accurate kinematics seem to be of higher importance for wear prediction than accurate loads. As discussed above, the ISO_FC,mod,nom_ models’ output kinematics deviated from the ones measured in the experiment by only a few mm/degrees but resulted in a threefold difference in wear rate. In contrast, the Stan_DC,mod,nom_ model’s loads deviated from the experiment by a factor of two for the internal moment and four for the anterior force, but still predicted wear rates accurately. The relatively large axial force is likely the main load driver for wear, while transverse load errors in DC models are less consequential. Reinforcing this deduction, other studies have also shown that variations in the joint kinematics e.g. in AP and IE directions (McEwen et al., 2005) had a larger impact on wear predictions than changes in the applied AP and IE loads (Lin et al., 2010; Pal et al., 2008).

Applying the Stan kinematics and associated CAMS-HIGH100 loads resulted in almost three times more wear than applying the ISO FC boundary conditions. While this is the first wear simulation study using the recently published Stan dataset, others have applied in vivo loads collected earlier from some of the same subjects with instrumented implants in FC mode. The reported wear rates, compared to the ISO FC BCs, exhibited large variability, going from comparable (Wang et al., 2019), slightly higher (Shu et al., 2021), to up to three times higher (Reinders et al., 2015). However, such comparisons between test standards may not necessarily yield the same results for other implant designs (Abdelgaied et al., 2022), prohibiting a general interpretation of these results. Yet, while the body of evidence is still small, these and this study’s results suggest that wear testing boundary conditions derived from in vivo measurements induce more wear than the standard ISO FC conditions.

A limitation of this study is that only one ultra-congruent implant design in one combination of component sizes and two sets of boundary conditions with different control methods were investigated. Further investigation of other implant designs and boundary conditions, e.g. the DC ISO standard and Stan’s other activities of daily living, should be considered to more comprehensively investigate the effects of BCs derived from in vivo measurements on wear testing outcomes. Furthermore, the FE models presented here did not model the variability in component positioning, loads, kinematics, and geometry, which is unavoidable on a knee simulator, and thus did not account for rare extreme motions and their possible impact on surface deformations and wear rates. Only the articulating surface was considered and only abrasive wear was modelled. The size of the contact patches was not measured experimentally, so no validation of the modelled contact area, which may have influenced predicted wear rates, was possible. Lastly, the PE material density and creep model were not obtained for the specific PE material investigated here. Notwithstanding these limitations, the experimental wear rates were accurately reproduced by the FE models, which were based on independent prior studies of PE mechanical properties and wear.

We recommend that future computational wear simulation studies not only use in vivo kinematics, but also consider multiple activities of daily living (Reinders et al., 2015; Shu et al., 2021) and incorporate uncertainty in their evaluation to account for the sensitivity of wear models to variations in contact mechanics. This could be achieved using available standardized BCs (Abdel-Jaber et al., 2016; ASTM International, 2017b; Bergmann et al., 2014; Dreyer et al., 2022b) complemented with a sensitivity analysis (Pal et al., 2008) or by modelling wear using data of multiple patients and trials (Fregly et al., 2012; Taylor et al., 2017). To make these rich datasets accessible for preclinical evaluation of implant wear, e.g. for different implant designs and patient specific factors, the validated WearPy software is available upon request at https://www.empa.ch/web/s304/wearpy.

## Supporting information

Verification & Validation

## Data Availability

The Stan dataset used as input in this study can be found at https://cams-knee.orthoload.com. The wear prediction algorithm can be requested at https://www.empa.ch/web/s304/wearpy. Data pertaining to the investigated implant, such as geometry or material data, is not available due to its confidential nature.

https://cams-knee.orthoload.com/data/

https://www.empa.ch/web/s304/wearpy

## Acknowledgements

We would like to thank Zimmer Biomet for providing access to the INNEX implant CAD files and PE material data, for providing material samples and implant components, and for running the physical test on the knee simulator.

