## Supplementary material for "Experimental and computational evaluation of knee implant wear and creep under in vivo and ISO boundary conditions": Verification & Validation

### Assessing the credibility of computational *WearPy* wear models per the ASME V&V 40 standard

#### Background

To apply the ASME V&V 40 [1] standard to our study, we propose a hypothetical scenario where WearPy is used in development of a new knee implant, which is based on an existing device with previous testing and satisfactory clinical history but different from the implant tested in the validation activities. The differences between the existing and the hypothetical new device are strictly geometric, with a change of the liner design. The material pairing (CoCr vs. PE), indications and size offerings are identical. The existing implant is the implant from our study that was used in the validation activities. For the wear evaluation of the new device, only displacement control boundary conditions are used. The goal is to identify the worst-case scenario, which will later be physically tested for regulatory approval.
This example is for illustration purpose; for any new study that would leverage the work presented in the current article, the context of use, question of interest, model risk, credibility goal and validation assessment would need to be reconsidered.
The gradations of credibility activities (“a”, “b”, “c”, “d”) follow the ASME V&V 40 standard.

#### 1. Context of use

Question of Interest: Does the hypothetical new total knee arthroplasty design provide sufficient resistance to wear of the polyethylene (PE) inlay under ISO 14243-1 [4] and activities of daily living [2] test conditions in displacement control?

Context of Use: WearPy is used to determine the amount of PE volumetric wear of the new design and identify the worst case condition (test and size). Bench testing will be performed on the identified worst-case.

#### 2. Model risk

2.1. **Model influence:** The model results will be supplemented with a physical test on the device of interest. Results will be compared to previous testing on a marketed device with satisfactory clinical history. Model influence: (a)

2.2. **Decision consequence:** An incorrect wear comparison between different configurations could lead to improper wear assessment and release of a product with insufficient wear resistance, which could lead to a revision of the knee implant. The decision consequence is (b).

Model risk: The model risk is determined to be LOW-MEDIUM

#### Credibility Target

With a LOW-MEDIUM model risk the credibility target is LOW-MEDIUM.

### 3. Model Credibility

#### 3.1. Verification

##### 3.1.1. Code verification

###### 3.1.1.1. Software Quality Assurance (SQA)

WearPy builds on Abaqus, an off-the-shelf software that is validated by the developer according to ISO 9001:2015 [5]. Abaqus was not validated internally with specific tests.

WearPy is a user-developed software for Abaqus. End-to-end tests of the WearPy pipeline with different input models (Abaqus Standard and Explicit, linear and quadratic element types, different geometries) were implemented to ensure that no software exceptions occurred . Very basic unit tests of single functions/classes were implemented. No code review by a third party was performed.

→ SQA Credibility: (a)

###### 3.1.1.2. Numerical Code Verification (NCV)

The implementation of the wear and creep material laws was checked by simulating simple pin-on-disk and uniaxial creep tests and comparing the results with the analytical solution. The predictions matched the analytical results within 3% and 1 % for wear and creep, respectively.

→ NCV credibility: (b)

##### 3.1.2. Calculation Verification

###### 3.1.2.1. Discretization Error

Several convergence analyses were performed as described below. Individually, all these discretization error sources were less than 3%.

Temporal convergence: The knee implant model of the ISO force-controlled condition was evaluated with 50, 100, 200, and 400 time steps per motion cycle. Convergence was achieved at 200 time steps with a <0.2% relative difference to 400 steps.

Maximum absolute surface deformation per wear analysis cycle increment ($n_{inc}$): A pin-on-disk test of a pin with a spherical tip (⌀5 mm) was simulated with quadratic tetrahedral mesh sizes of 0.1 and 0.05mm, normal forces of 100 and 10N, and $n_{inc}$ of 0.02 and 0.01. With an $n_{inc}$ of 0.01, the wear volume error compared to the analytical solution was <3% for all combinations of the other parameters.

Mesh size: The mesh size was converged for one preliminary knee implant model of the ISO force-controlled condition with respect to wear. For the inlay, quadratic tetrahedral element sizes of 2mm, 1mm and 0.5mm were evaluated. For the femoral component, mesh sizes of 2mm, 1mm, 0.5mm, and 0.25mm were evaluated. Convergence was achieved with an inlay element size of 1mm at <3% difference to a larger mesh. Differences between choosing a femoral element size of 0.5mm or 0.25mm were negligible and 0.5mm was chosen.

→ Discretization error credibility: (c)

###### 3.1.2.2. Numerical Solver Error

No numerical solver error credibility was performed.

→ Numerical Solver Error credibility: (a)

###### 3.1.2.3. Use Error

Key inputs and outputs were verified by the practitioner, but not by internal peer review.

→ Use error credibility: (b)

#### 3.2. Validation

##### 3.2.1. Computational Model

###### 3.2.1.1. Model Form

The effect of applying force- vs displacement-controlled boundary conditions was investigated and found to be substantial (see main publication). For this reason, the COU model is intended to use displacement-controlled boundary conditions.

The pin-on-disk model was used to investigate the effect of various penalty contact formulations. While the default linear stiffness resulted in an error of 9% compared to the analytical solution, reduction of the stiffness by a factor of 10 or the choice of the standard non-linear stiffness reduced the error to 2%, and a nonlinear stiffness reduced by a factor of 100 reduced the error to 0.4%.

The effect of activating tangential contact damping instead of having no damping was checked for a preliminary force-controlled ISO model. A fixed damping value of 0.0001 and automatic damping scaled by 0.001 and 0.0001 were investigated. For automatic damping scaled by 0.001, up to 2mm difference in anterior-posterior translation was found. For the other two smaller damping values, differences did not exceed 0.3mm.

The quantitative influence of other model form assumptions such as constitutive, wear, or creep models, solver solution schemes, exact type of quadratic element type, etc. was not quantitatively investigated.

→ Model form credibility: (b)

###### 3.2.1.2. Model Inputs

###### 3.2.1.2.1. Quantification of Sensitivities

The influence of deviations of several mm/degree in displacement control was investigated (see main publication). Other sensitivities to input loads and kinematics, coefficient of friction, posterior tibial slope, etc. were not investigated.

→ Quantification of sensitivities credibility: (b)

###### 3.2.1.2.2. Quantification of Uncertainties

Uncertainties were not identified.
→ Quantification of Uncertainties credibility: (a)

##### 3.2.2. Comparator

The physical tests of Innex implants on the AMTI knee simulator under ISO 14243-1 and Stan conditions (see main publication) serve as the comparator for this study.
The main output was quantitative PE wear volume. Secondary, surface deformation due to wear and creep damage was visually evaluated.

###### 3.2.2.1. Test Samples

###### 3.2.2.1.1. Quantity of Test Samples

Three test samples per condition were used for each condition as per the ISO 14243-1 standard but no power analysis was performed.
→Quantity of test samples credibility: (b)

3.2.2.1.2. **Range of Characteristics of Test Samples**

A single implant size was tested.

→ Range of characteristics of test samples credibility: (a)

3.2.2.1.3. **Measurements of Test Samples**

All samples were production parts and dimensions were controlled to be within the production tolerance. Material properties for the polyethylene and metal components were within the material specifications. Surface roughness of the femoral component, tibial inlay, and tibial baseplate were measured before and after the test in multiple locations.

→Measurement of test samples credibility: (c)

3.2.2.1.4. **Uncertainty of Test Sample Measurements**

Samples were characterized with calibrated equipment, so that uncertainty analysis incorporated instrument accuracy only.

→uncertainty of test sample measurements credibility: (b)

**3.2.2.2. Test Conditions**

**3.2.2.2.1. Quantity of Test Conditions**

Two conditions were examined on the knee simulator (ISO 14243-1 and Stan)

→Quantity of test conditions credibility: (b)

**3.2.2.2.2. Range of Test Conditions**

Test conditions across a range of expected normal conditions (regular ISO 14243-1 and representative Stan conditions in the knee test) were examined.

→Range of test conditions credibility: (b)

**3.2.2.2.3. Measurements of Test Conditions**

Lubricant protein content, posterior tibial slope, loads and kinematics, lubricant temperature on the machine were measured.

→Measurement of test conditions credibility: (c)

**3.2.2.2.4. Uncertainty of Test Condition Measurements**

Calibrated equipment was used to perform key test condition measurements, but no assessment of intra- or inter-operator variability was performed.

→Uncertainty of test condition measurements credibility: (b)

**3.2.3. Assessment**

**3.2.3.1. Equivalency of Input Parameters**

As the model reproduced the physical test, all inputs were equivalent (same implant type and size, same loading and kinematics, same set up).

→Equivalency of input parameters credibility: (c)

**3.2.3.2. Output Comparison**

**3.2.3.2.1. Quantity**

Wear volume and knee implant deformation were evaluated.

→Quantity credibility: (b)

###### 3.2.3.2.2. Equivalency of Output Parameters

Types of outputs (wear volume, surface damage) were equivalent.

→Equivalency of output parameters credibility: (c)

###### 3.2.3.2.3. Rigor of Output Comparison

Wear volume was compared quantitatively (no model uncertainty, sample variability) and knee implant deformation was compared visually (model contour plots, sample 3D scan).

There was no consideration of uncertainty.

→ Rigor of output comparison Credibility: (b)

###### 3.2.3.2.4. Agreement of Output Comparison

In displacement control, good agreement was obtained for volumetric wear, with the model output falling within the ranges of experimental outputs. Visually good agreement of damage patters was obtained.

→Agreement of output comparison Credibility: (b)

##### 3.3. Applicability of the Validation Activities to the Context of Use (COU)

Note: The applicability is based on the hypothetical scenario/context of use outlined at the beginning of this document. Any study leveraging this V&V should critically consider the applicability for the specific COU.

The COU encompasses all of the validation points.

###### 3.3.1. Relevance of the QOIs

Both the validation activities and the COU are focused on volumetric wear and surface damage as the QOI.

→Relevance of the quantities of interest credibility: (c)

###### 3.3.2. Relevance of the Validation Activities to the COU

The geometries, materials, and boundary conditions for the validation activities are within the range of associated parameters for the COU.

The hypothetical device of interest has small geometric differences to the implant used in the validation activities, but it is also a knee implant of similar type and size, for identical indications and made of identical material pairing (CoCr vs. PE).

The COU and validation activities implement an identical wear test.

→Relevance of the validation activities to the COU credibility: (d)

#### 4. Summary

The gradation of all credibility factors is summarized in figure S1, following the example proposed by the recent FDA draft guidance [3].

The credibility of four factors ("Software Quality Assurance", "Numerical Solver Error", "Quantification of Uncertainties of the Computational Model", and "Range of Characteristics of Comparator Test Samples") were lower than the credibility target. Given the credibility of the other factors, all of which meet or exceed the low-medium credibility target, the overall credibility of the modeling approach can be considered sufficient.

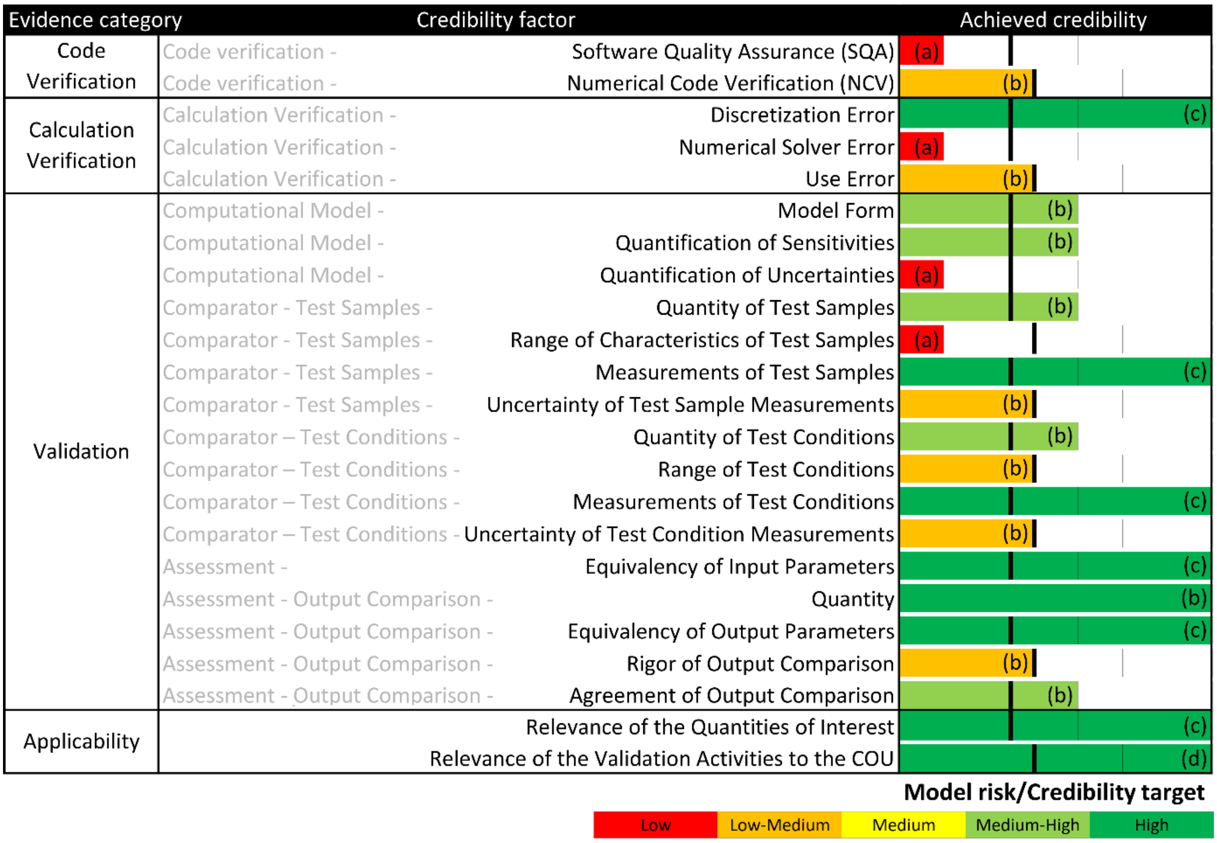


Figure S1. Achieved credibility for all credibility factors, inspired by the example proposed in the recent FDA draft guidance document [3]. The color coding and length of the horizontal bars indicate the achieved level of credibility, and the vertical black line segments indicate the model risk/credibility target. Mapping the variable 2 to 4 level gradation from the ASME V&V40 [1] to a five-level gradation scheme from the FDA required an adaptation of the model risk/credibility target line to each credibility factor.
